## Supplementary material for "Acceptability and feasibility of tests for infection, serological testing and photography to define need for interventions against trachoma": S1 - Topic Guides

### Topic Guide: Focus Group Discussion (FGD)

| **Research question: Among communities in trachoma endemic or formerly endemic areas, what is the acceptability of blood spot collection, eye swab collection, and conjunctival photography for trachoma surveillance purposes?** | |
| --- | --- |
| **Key areas of investigation** | **Example topics and questions (adjusted by context, group and iterative analysis)** |
| **Warm up** | - Introductions within group - Ice-breaker and ground rules for confidentiality and respect within the FGD - Brief explanation of trachoma and the purpose of the FGD |
| **Clinical grading of TF** | - Show image of someone having their eyes examined for trachoma - What is your experience of having a clinical exam for trachoma? Have you ever had a clinical exam for trachoma? Have your children and/or the children in your care ever had a clinical exam for trachoma? - Explain the exam: The main purpose of this activity is to determine the amount of trachoma in a community. A trained grader comes to the community and examines children's eyes for trachoma. In this exam, the eyelid is flipped in order to look for signs of a current infection with the germ that causes trachoma. If a person has the signs, they are given treatment to clear the infection. The grader will also check for advanced stages of the disease, and a referral for surgery will be given if necessary. The results are used to decide whether the whole community should receive treatment or not. If treatment is needed, this is given at a later date. - What do you think about the exam process? What do you think are some advantages of a clinical exam? What do you think are some disadvantages? - If someone came to your community and examined your child's eyes for trachoma in order to determine the amount of trachoma in your community, would you want to participate? Why or why not? |
| **Conjunctival photography** | - Show image of someone having their conjunctiva photographed with a smartphone - What is your experience of having a photo of your eyelid taken for trachoma? Have you ever had a photo of your eyelid taken? Have your children and/or the children in your care ever had a photo of their eyelid taken? - Explain the exam: The main purpose of this activity is to determine the amount of trachoma in a community. A trained photographer comes to the community and uses a camera to take a photo of the inside of your eyelid. This photo is then sent electronically to a person trained to look for signs of the germ that causes trachoma. This person might be in another country. In this type of exam, you would not know if you individually have the pathogen. The results are used to decide whether the whole community should receive treatment or not. If treatment is needed, this is given at a later date. - What do you think about the exam process? What do you think are some the advantages of an eyelid photo being taken? What do you think are some disadvantages? - If someone came to your community and took a photo of your eyelid in order to determine the amount of trachoma in your community, would you want to participate? Why or why not? Would you want your children to participate? Why or why not? |
| **Eye swabs** | - Show image of someone having their eyes swabbed for trachoma - What is your experience of having an eye swab for trachoma? Have you ever had an eye swab? Have your children and/or the children in your care ever had an eye swab? - Explain the exam: The main purpose of this activity is to determine the amount of trachoma in a community. A trained individual comes to the community and uses a clean cotton swab to take a sample from the inside of the eyelid. The swab is then sent off to a lab to test for the germ that causes trachoma. This lab might be in another country. In this type of exam, you would not know if you individually have the pathogen. The results would be used to decide whether the whole community should receive treatment or not. If treatment is needed, this would be given at a later date. - What do you think about the exam process? What do you think are some the advantages of an eye swab? What do you think are some disadvantages? - If someone came to your community and swabbed your eyes for trachoma in order to determine the amount of trachoma in your community, would you want to participate? Why or why not? Would you want your children to participate? Why or why not? |
| **Blood spots** | - Show image of someone having their finger pricked/a dried blood spot being taken - What is your experience of having a blood spot taken for trachoma? Have you ever had a blood spot taken for any other disease (ex: malaria)? Have your children and/or the children in your care ever had a blood spot taken? - Explain the exam: The main purpose of this activity is to determine the amount of trachoma in a community. A trained individual comes to the community and uses a clean needle to prick your finger. A small amount of blood is then collected onto filter paper. The filter paper is then sent off to a lab to test for evidence in the blood of a previous infection with the germ that causes trachoma. This lab might be in another country. In this type of exam, you would not know if you individually have the pathogen. The results would be used to decide whether the whole community should receive treatment or not. If treatment is needed, this would be given at a later date. - What do you think about the exam process? What do you think are some the advantages of a blood spot? What do you think are some disadvantages? - If someone came to your community and took a blood spot in order to determine the amount of trachoma in your community, would you want to participate? Why or why not? Would you want your children to participate? Why or why not? |
| **Preference between methods** | - Show cards with images of each method and ask the group to order the methods from most-preferred to least-preferred. - Are there any methods to check for trachoma in your community that you would not want to participate in? Why or why not? Are there any methods that you would not want your child/a child in your care to participate in? Why or why not? - If there were a lot of people going blind in your community, would you be more likely to participate and/or have your children participate? What about if there were not a lot of people going blind in your community?   The **clinical exam** sign is an accurate indicator of the germ that causes trachoma in the individual in areas with a lot of trachoma, but as trachoma in a community decreases, the sign can become less accurate. The **photography** exam sign is an accurate indicator of the germ that causes trachoma in the individual in areas with a lot of trachoma, but as trachoma in a community decreases, the sign doesn't tell us as much. **Eye swab testing** is accurate for identifying the germ in the individual, even in areas with very low levels of trachoma. The **blood spot test** tells us about the current and previous presence in the community of the germ that causes trachoma in the community.   - How would you feel if any of these methods were used on a routine basis to monitor the level of trachoma in your community over time (say, every couple of years or so)? Would you have a preference for which type of test was done? Which test, and why? (Or if no preference, why not?) |
| **Wrap-up** | Thank participants for their time and provide space for further discussion of relevant topics. |

### Topic Guide: In-depth Interviews (IDI)

| **Research question: 2. Among public health practitioners in trachoma endemic or formerly endemic areas, what is the perception of the feasibility of blood spot collection, eye swab collection, and conjunctival photography for trachoma surveillance purposes?** | |
| --- | --- |
| **Key areas of investigation** | **Example topics and questions (adjusted by context, group and iterative analysis)** |
| **Warm up** | - Explanation of the purpose of the IDI |
| **General information /experience** | - Which organization are you associated with and what is your role within this organization? - Could you tell me about the role of this organization in the surveillance of trachoma? - Could you tell me about the role of this organization in the surveillance of other diseases (such as HIV, Malaria, other NTDs)? |
| **Current diagnostic/surveillance methods** | - How much trachoma is in your area? - Can you tell me how trachoma is currently diagnosed in your area? - What are the main challenges for diagnosing trachoma in your area? - Which sample types are currently used for the surveillance of trachoma in your area? Why are these sample types used? Are there sample types you think are better but don't currently use? - Can you tell me about other disease programmes in your area that use infection testing as a part of routine surveillance efforts? - Can you tell me about other disease programmes in your area that test blood samples as a part of routine surveillance efforts? |
| **Surveillance (long-term)** | - What is the long-term plan for the surveillance of trachoma, i.e. once elimination has been achieved? Why? What is the rationale for this plan? How feasible do you think it is? - What do you see as the main challenges in the surveillance of trachoma in your area? |
| **Clinical examination** | - What do you think about using examination of the eyelid to check for the presence of trachoma (TF grading)? - Why? Do you and your organization consider this practice acceptable? What are the advantages to using this procedure? Are there any drawbacks to using this procedure? |
| **Conjunctival photography** | - What do you think about taking a picture of the eye to check for presence of trachoma? - What are your thoughts on the feasibility of taking pictures as part of trachoma surveys? For example, what would expect the impact to be on fieldwork time? What would you expect the impact to be on cost? What would you expect the impact to be on obtaining data results? Are there any other concerns you can think of? - What do you think would be some advantages of taking photos of people's eyelids? What might be some disadvantages? |
| **Eye swabs/ *Ct* infection testing** | - What do you think about using a swab of the eyelid to be able to test for the infection causing trachoma? - What are your thoughts on the feasibility of collecting eye swabs as part of trachoma surveys? For example, what would expect the impact to be on fieldwork time? What would you expect the impact to be on cost? What would you expect the impact to be on obtaining data results? Are there any other concerns you can think of? - What are your thoughts on the feasibility of testing eye swabs for *Ct*? Do you think this could be done in your area? Why or why not? Do you think your area has the appropriate lab capacity to do this? Why or why not? - Do you have experience w/ tests requiring a cold chain? Do you see any barriers to a test requiring a cold chain? Why or why not? - What do you think would be some other advantages of collecting/testing eye swabs? What might be some disadvantages? |
| **Blood spots/serology** | - What do you think about using a dried blood spot to check for current or past transmission of *Ct* at the community level? - What are your thoughts on the feasibility of collecting blood as part of trachoma surveys? For example, what would expect the impact to be on fieldwork time? What would you expect the impact to be on cost? What would you expect the impact to be on obtaining data results? Are there any other concerns you can think of? - What are your thoughts on the feasibility of testing blood for the presence of trachoma? Do you think this testing could be done in your area? Why or why not Do you think your area has the appropriate lab capacity to do this? Why or why not? - What do you think would be some advantages of collecting/testing dried blood spots? What might be some disadvantages? |
| **Wrap-up** | Thank participants for their time and provide space for further discussion of relevant topics. |
