## Supplementary material for "Acceptability and feasibility of tests for infection, serological testing and photography to define need for interventions against trachoma": S2 - a priori Codes

### Tanzania Alternative Indicators

#### Codes

| Name | Description |
|---|---|
| Blood advantages | Advantages to swabs (compared to other test types) |
| Blood disadvantages | Disadvantages of blood spots (compared to other test types) |
| Blood experience | Participant has experience with blood spot |
| Blood would not participate | If participant would not participate in blood spot |
| Blood would participate | If participant would participate in blood spot |
| Photo advantages | Advantages to photography (compared to other test types) |
| Photo disadvantages | Disadvantages of photography (compared to other test types) |
| Photo experience | Whether or not participants have experience with conj. photography. (Positive/negative experiences will be coded under advantages/disadvantages.) |
| Photo would not participate | If participant would not participate in photography |
| Photo would participate | If participant would participate in photography |
| Swab advantages | Advantages to swabs (compared to other test types) |
| Swab disadvantages | Disadvantages of swabs (compared to other test types) |
| Swab experience | Participant has experience with eye swab |

| Name | Description |
| --- | --- |
| Swab would not participate | If participant would not participate in eye swab |
| Swab would participate | If participant would participate in eye swab |
| test ranking | Direct comparison of test types to each other (sentiments like "better","worse", "I would prefer", etc.) |
| TF advantages | Advantages to TF grading (compared to other test types) |
| TF disadvantages | Disadvantages of TF grading (compared to other test types) |
| TF experience | Whether or not participants have experience with TF grading. (Positive/negative experiences will be coded under advantages/disadvantages.) |
| TF would not participate | If participant would not participate in TF grading. |
| TF would participate | If participant would participate in TF grading. |
