## Supplementary material for "Acceptability and feasibility of tests for infection, serological testing and photography to define need for interventions against trachoma": S5 - Knowledge Gaps

| Category | Key questions |
|---|---|
| Setting/interpreting meaningful thresholds | What is the threshold for infection? How does this vary in different settings? Will the indicator be presence/absence of *Chlamydia trachomatis (Ct)* , or will typing occur to determine serotype? |
| | What is the threshold for serology? How does this vary in different settings? How do age dynamics affect thresholds/cutoffs? |
| | What is the average change in trachomatous inflammation--follicular (TF) prevalence from trachoma impact survey (TIS) to trachoma surveillance survey (TSS)? |
| | Is the 5% prevalence threshold for TF in children aged 1-9 years adequate to ensure that trachoma will not spread in the community? |
| | How can we trust/validate any thresholds? |
| | Can community-level (vs. other-level) thresholds be set? How to understand infection results at community |
| Test characterstics (general) | What is the sensitivity/specificity/positive predictive value (PPV)/negative predictive value (NPV) of each test? |
| Test characterstics (photography) | How accurate is human grading compared to grading via photography with artificial intelligence (AI)? Does this differ by setting? |
| | How to interpret any increased sensitivity of photography compared to field grading of TF? Should we consider follicles outside the inclusion zone or <0.5mm? |
| Survey design/screening protocols | Can we develop models for integrating other indicators (including other clinical signs) into trachoma screening to accurately estimate infection? |
| | Can other indicators be used to confirm a TF diagnosis? |
| | Has the current method of pooling been validated? What's the best number of samples to pool to bring down cost/time of analysis without a loss in accuracy? |
| | What infection testing platform would be best: multiplex PCR or could a point-of-care (POC) test be useful? |
| | What's the best age group to test? |
| | How many people need to be tested? Does this differ by TF prevalence setting? |
| | What testing protocol produces best sensitivity/specificity/PPV/NPV? |
| Interpreting serological data | How to interpret different sero- and sub-types of Chlamydia? (ex: ocular vs. STI) |
| | How to interpret role of seroreversion? |
| | What is the relationship between seroconversion due to ocular exposure vs. other routes? |
| | What is the load or frequency of infection required to cause seroconversion? |
| | How do we test for/interpret previous exposure? |
| Clinical signs/progression of disease | What is the duration of clinical signs in areas with formerly high levels of trachoma? |
| | What is the temporal variability of infection (i.e. how much does exact timing of swab collection matter vs. TF, which is positive for longer and so timing matters less)? |
| | How does infection relate to fine-graded TF in an individual? (All we have is pooled/community level data.) |

| Category | Key questions |
| --- | --- |
| | Why is there a lag between clearance of infection and resolution of TF? |
| | What is the progression from infection to TF? |
| | In settings with a formerly high level of trachoma, are clinical signs indicative of scarring? |
| | In surveillance settings, are clinical signs sufficiently rapid and sensitive to detect infections that will lead to later scarring/trachomatous trichiasis (TT)? |
| | What evidence supports the number, size, and position of follicles in the definition of TF? |
| | Should resolving follicles (from children treated with azithromycin) be included in the calculation of TF prevalence? |
| | How to differentiate dead follicles from active ones? |
| | What's the longitudinal trajectory of infection? |
| | What is the relationship between seropositivity and continued scarring progression once elimination thresholds have been reached? |
| Integration within existing systems | How to integrate infection testing into ongoing surveys done by national programmes? |
| | Can TF grading be incorporated into primary care/routine child health care/school-based programs to be used effectively as post-elimination surveillance to detect potential recrudescence |
| | How can infection testing be integrated with other neglected tropical disease (NTD) testing? |
| | How to integrate photography with other data collection platforms? |
| | Can we certify graders using Tropical Data (TD) system for all eye health care workers to routinely conduct TF grading? |
| Feasibility/acceptability issues | What are the ethical considerations of photography (concerns about anonymity) and will this increase training time/survey time and therefore cost? |
| | What are the costs compared to other indicators? |
| | Are photos acceptable to communities? |
| | How can AI be used (with photography) and what is the cost of this? |
| | How can photography be used at scale? |
| | Is infection testing acceptable to communities? |
| | What's the cost at-scale and how can the cost be reduced by using regional testing centers? |
| | How to handle increased logistics needs for infection testing? |
| | What's the additional costs associated with sample management vs. the sampling itself? |
| | What is the community acceptability of serology? |
| | How to ensure adequate funding? |
| | How to ensure trust with communities that samples will be used only for trachoma testing? |

| Category | Key questions |
|---|---|
| | What will the cost reduction be by not doing mass drug administration based (only) on TF results? |
| | What are the actual relative costs of TF, photography, eye swabs, serology? What does "increased costs" actually signify? |
| | How much, for each indicator, is there capacity in-country, and how much are countries reliant on external partners? |
| Mitigating training/lab quality issues | How to mitigate the loss of skill of TF grading in low-prevalence/post-elimination settings? |
| | How can we standardize training and grading? |
| | How to ensure quality photos? |
| | How to build lab capacity in country or regional settings? |
| | How to train people adequately? |
| | Which camera equipment best? |
| | How to upload/share quality photos? |
| | How to ensure consistency in photography use/specifications/training, etc.? |
| | How to monitor for lab contamination? |
| | How to ensure safety of community members/training of swab-takers? |
| | How to ensure adequate lab capacity/training to ensure quality samples/standardized analysis? |
| | How to mitigate the difficulty and inherent subjectivity of grading/interpreting results in low-prev settings? |
| | How long does it take on average from sample collection to results generation across multiple settings? |
| | What's the stability of swabs in different settings and time to arrival to lab? |
| other | Is DNA detection good enough, or should we be looking for viable organisms? |
| | Why is there recrudescence of TF in the absence of infection/other signs? |
| | How to ensure development/roll out of a rapid diagnostic test? |
